# Differential expression of selected microRNA and putative target genes in peripheral blood cells as early markers of severe forms of dengue

**DOI:** 10.1101/19002725

**Authors:** Harsha Hapugaswatta, Pubudu Amarasena, Ranjan Premaratna, Kapila N. Seneviratne, Nimanthi Jayathilaka

**Affiliations:** Department of Chemistry, Faculty of Science, University of Kelaniya, Kelaniya, Sri Lanka; North Colombo Teaching Hospital, Ragama, Sri Lanka; Department of Medicine, Faculty of Medicine, University of Kelaniya, Kelaniya, Sri Lanka

**Keywords:** Dengue, Dengue hemorrhagic fever, microRNA, acute dengue biomarkers

## Abstract

**Background:** Dengue presents a wide clinical spectrum including asymptomatic dengue fever (DF) or severe forms, such as dengue hemorrhagic fever (DHF) and dengue shock syndrome (DSS). Early symptoms of DHF are similar to those of non-life-threatening DF. Severe symptoms manifest after 3-5 days of fever, which can be life threatening due to lack of proper medications and inability to distinguish severe cases during the early stages. Early prediction of severe dengue in patients with no warning signs who may later develop severe infection is very important for proper disease management to alleviate DHF related complications and mortality. Due to the role in post-transcriptional regulation of gene expression and remarkable stability of microRNA, altered expression of microRNA was evaluated to explore clinically relevant biomarkers.

**Methodology/Principal findings:** The relative expression of microRNA hsa-let-7e, hsa-miR-30b-5p, hsa-miR-30e-3p, hsa-miR-33a, and hsa-miR-150-5p and several putative target genes in peripheral blood cells (PBC) collected from 20 DF and 20 DHF positive patients within four days from fever onset was evaluated by qRT-PCR. hsa-miR-150-5p showed significant (P<0.05) up regulation in PBC of DHF patients compared to DF patients during the acute phase of infection. Expression of enhancer of zeste homolog 2 (EZH2) was significantly (P<0.05) down regulated indicating that genes involved in epigenetic regulation are also differentially expressed in DHF patients during the early stage of infection.

**Conclusions/Significance:** Differential expression of microRNA miR-150-5p and the putative target gene EZH2 may serve as reliable biomarkers of disease severity during early stages of dengue infection.

**Author summary:** Severe dengue cannot be distinguished from dengue fever during the early stages of infection based on the clinical symptoms. A diagnosis is only made after the patient is presented with severe manifestations such as plasma leakage and hemorrhage. During a dengue outbreak, this leads to high occupancy of hospital beds. However, only a small percentage of patients present with severe symptoms and the others do not require medical care at a hospital. Therefore, early prognosis of severe manifestations could reduce dengue related mortality by identifying the patients who will benefit from hospitalization and early intervention. We demonstrate that severe dengue in Sri Lankan patients is associated with increased expression of miRNA miR150 and decreased expression of EZH2 during the early stages of infection when none of the patients showed symptoms of developing severe manifestations at later stages of infection.

## Introduction

More than half of the world population is at risk of contracting dengue, a mosquito-borne viral infection with approximately 50 to 100 million cases reported each year in many tropical countries including Sri Lanka [1]. In 2018, 51659 suspected dengue cases have been reported to the Epidemiology Unit of Sri Lanka from all over the island making it the most important tropical disease posing a significant public health threat. Dengue fever (DF) is characterized by headache, retro orbital pain, body pain, nausea, vomiting, joint pains and weakness [2]. Severe manifestations of dengue, dengue hemorrhagic fever (DHF) and dengue shock syndrome (DSS) also show similar symptoms during the early stages of infection. After 3-5 days from fever onset, DHF patients manifest plasma leakage, elevated hematocrit and pleural effusions. Humans possessing pre-infection antibody, passively acquired or derived from heterotypic infection are susceptible to DHF [3]. There are four dengue virus serotypes each of which is capable of infection [4]. DENV1 and DENV2 have been reported as prevalent serotypes with a similar number of reported infections [5]. All four virus serotypes are present in Sri Lanka. DENV2 and DENV3 are the common serotypes reported in many parts of Sri Lanka, with DENV3 reported to be responsible for many of the infections that progress to DHF [6]. Inability to distinguish severe dengue from DF during the early stages of infection makes this disease life threatening.

There are no proper medications or vaccines available for prevention and treatment of dengue [7]. The first ever dengue vaccination program was launched in 2016 in Philipines with complications rising from increased risk of severe dengue in particular circumstances [1]. Therefore, dengue prevention is largely limited to vector control. Several research groups have reported potential small molecules for drug intervention of DF [8-10]. In addition, there is ongoing research on antiviral activity of phytochemicals as well as currently available antiviral drugs against dengue viruses. However, these phytochemicals and drugs have not been approved for the management of dengue infections [11,12]. Diagnostic tests based on PCR or serological testing for dengue do not distinguish DHF from DF and disease severity is determined after the patient is presented with severe symptoms [13]. However, if diagnosed early, effective disease management only involves hospital care and hydration to mitigate complications from severe dengue [14]. Since there are no early clinical tests approved for prognosis of severe dengue, most patients suspected of having contracted dengue are hospitalized for disease management resulting in high healthcare costs, though only a small fraction of DF patients develop DHF and DSS. Therefore, identification of markers for the early detection of DHF could effectively cut down dengue related mortality as well as cost of healthcare during dengue outbreaks.

Several studies comparing the transcriptomes of DF and DHF patients have reported increased levels of cytokines, TNF-α, IL-2, IL-6, IL-10, IL-12 and IFN-γ involved in host immune responses in DHF patients [15-19]. Genetic varients in MICB and PLCE1 has been found to be associated with severe dengue [20]. However, these studies have not determined whether these markers serve as differential markers during the early stage of infection when severe manifestations of dengue cannot be distinguished based on the clinical symptoms.

MicroRNA molecules are small non coding RNA molecules with the length around 20-22 nucleotides. Due to their role in regulating the expression of putative target genes, microRNA are important for function of human cells. Therefore, microRNA show characteristic expression in different diseases. Since microRNAs are also remarkably stable in blood, they are under investigation as potential biomarkers. Recent studies have reported differential expression of microRNA in dengue patients and infected cultured cells [21,22]. hsa-let-7e, hsa-miR-30b-5p and hsa-miR-33a have been found to differentially express in dengue virus infected cultured cells [23]. Similarly, miR-30e-3p also express differentially in dengue infected cultured cells [24]. miR-150 expression is upregulated in DENV infected cultured cells within few hours of the infection [25]. Expression of miR150 is also found to be significantly upregulated in DHF patients compared to DF patients [26]. However, the expression level of miR-150 during the early stages of dengue infection, when prediction of disease outcome is not possible, has not been conducted in patient samples to determine the potential to serve as an early indicator of progression to severe dengue. Therefore, we evaluated the relative expression of microRNAs hsa-let-7e, hsa-miR-30b-5p, hsa-miR-30e-3p, hsa-miR-33a and hsa-miR-150-5p against the geometric mean of hsa-miR-16-5p and hsa-miR-103a-3p as reference genes and their putative target genes in peripheral blood cells from blood samples collected from dengue positive patients within four days of fever onset by qRT-PCR.

let-7e and hsa-miR-150-5p have been connected to regulation of the enhancer of zeste homolog 2 (EZH2) expression [27]. In addition, miR-30b-5p has been shown to regulate DNA methyl transferase 3 alpha (DNMT3A) expression that can be used as a potential biomarker and a therapeutic target for cancer [28]. DNMT3A has been found to regulate herpes simplex virus-1 propagation [29]. miR33a regulates the expression of receptor interacting protein 140 (RIP140) and ATP-binding cassette transporter subfamily A member 1 (ABCA1) involved in lipid metabolism and transportation, thus, play a role in viral invasion. miR-33a directly down regulates the expression of ABCA1 in cholesterol metabolism pathway and also contributes to the regulation of cholesterol and fatty acid homeostasis by targeting RIP-140 [30]. There is also evidence that DENV infection leads to an autophagy-dependent processing of lipid droplets and triglycerides to release free fatty acids which increases the cellular *β*-oxidation that generates ATP. These processes are required for efficient DENV replication [31]. ABCA1 expression inhibits Hepatitis C virus cell entry, acting on virus-host cell fusion [32]. Therefore, the relative expression of these putative target genes of above microRNA, EZH2, ABCA1, DNMT3A, and RIP140 were evaluated against GAPDH as the reference gene for their potential to serve as markers of early prognosis of severe dengue.

## Methods

### Ethics statement

Ethical clearance for patient sample collection was obtained from the Ethics Review Committee of the Faculty of Medicine, University of Kelaniya, Kelaniya, Sri Lanka (Reference number-P/119/07/2015). Written patient consent was obtained prior to the sample collection. The patient consent form was approved by the same ethics review committee.

### Sample collection and processing

Patients presented with clinical symptoms of dengue viral infection according to WHO Dengue case classification (fever, with two of the following criteria: headache, retro-orbital pain, myalgia, arthralgia, rash, hemorrhagic manifestations with no plasma leakage, and following laboratory findings: leucopenia, thrombocytopenia and rising haematocrit with no evidence of plasma loss) within 4 days from fever onset who tested positive for onsite NS1 rapid test (SD Bio) were recruited for the study from the North Colombo Teaching Hospital, Sri Lanka with informed consent. Patients who later develop DHF were determined according to the WHO guidelines; Fever and Hemorrhagic manifestation (positive tourniquet test) with evidence of plasma leakage (portable bedside ultrasonogram), spontaneous bleeding, circulatory failure, profound shock with undetectable BP and pulse, thrombocytopenia < 100 000 cells / mm^3^, and HCT rise > 20% [2]. 2.5 ml EDTA blood samples were collected from patients within 4 days from fever onset and transported and processed at 4 °C within 2 hrs from sample collection. Isolated peripheral blood cells and saliva samples were stored at - 80 °C until sample analysis.

### Quantitative Real-time PCR

MicroRNA primers were designed based on the miRbase database and gene specific human primers against EZH2, DNMT3A, RIP140, ABCA1 and the reference gene, GAPDH were mined from previously published literature (Tables S**1** and S**2**). Primers for microRNA target genes, were designed using NCBI primer picker to span exon-exon junctions. Total RNA was isolated from peripheral blood cells using miRNeasy Serum/Plasma kit (Qiagen) according to the manufacturer’s instructions. cDNA was synthesized using the miScript II RT Kit with Hiflex buffer for polyadenylation of microRNA according to product manual (Qiagen). Expression of both microRNA and mRNA target genes was quantified using miScript SYBR Green PCR kit according to product manual (Qiagen). Each reaction was carried out in triplicates in 20 μL reaction volume using StepOne Real-Time PCR System (Applied Bio Systems). The efficiency of amplification was above 80 % based on the standard curve analysis. No-template reactions and melting curve analysis was used confirm specificity of target amplification.

### Statistical Analysis

q-q plots and Shapiro-Wilk test were used to determine normality at a 95% confidence interval. For the Shapiro-Wilk test P>0.05 was determined as normal distribution. The fold change of microRNA and mRNA expression was calculated using the equation 2^−ΔΔCq^. A difference in expression based on log base 2, less than 0.5 was considered as downregulation and above 1.5 was considered as upregulation between DF and DHF cases. Statistical significance for differentially expressed targets were determined using independent t-test based on SEM of ΔCq. P < 0.05 was considered statistically significantly different. Logistic regression analysis for odds ratio, receiver operator characteristics, area under curve, specificity and sensitivity was determined using IBM SPSS Statistics, 2013 version at a 95% confidence interval. Sample sizes were calculated using G*Power 3.1.9.2 software at 95% confidence interval with a power of 80% for normally distributed samples using parametric test and skewed distributions using nonparametric test.

## Results

### Differential expression analysis of selected microRNA in dengue patients within four days from fever onset

Expression of hsa-let-7e, hsa-miR-30b-5p, hsa-miR-30e-3p, hsa-miR-33a, and hsa-miR-150-5p in patients suspected of having dengue based on clinical symptoms and positive result for NS1 rapid test was analyzed within four days from fever onset. The data were analyzed against the geometric mean of hsa-miR103a-3p and hsa-miR-16-5p reported to have stable expression in peripheral blood cells during infections. Patients were classified as dengue fever (DF) based on the clinical symptoms. Patients who later developed DHF were classified based on evidence of plasma leakage (pleural effusions and ascites) detected using a portable ultrasonogram. Twenty DF and 20 DHF samples were analyzed. Among them, most of the subjects were male (77%), with a median age of 30 (18-60) while the female subjects had a median age of 24 (19-60) years. At enrollment, there were no statistically significant differences in median laboratory clinical parameters including platelet count, hematocrit level, AST and ALT levels in patients who later developed DHF compared with DF (Table 1). Circulating AST and ALT levels were not significantly different between these two groups throughout the course of infection (Table S3). Therefore, DF patients could not be distinguished from the DHF patients during the acute phase of infection based on the clinical characteristics.

**Table 1.** Clinical characteristics of dengue patients recruited for microRNA and target gene expressions analysis at admission (median± MAD)

|  | DF patients<br>(n=20) | DHF patients<br>(n=20) | P value<br>CI, 95% |
| --- | --- | --- | --- |
| Gender (Male%/Female%) | 70/30 | 85/15 |  |
| Age | 30 (18-60) | 27 (19-60) |  |
| Platelet (x1000 /mm<sup>3</sup>) | 119.00±39.00 | 125.00±24.00 | 0.66 |
| Hematocrit (%) | 40.00±3.15 | 39.70±2.20 | 0.57 |
| Hemoglobin (g/dL) | 14.00±1.05 | 13.00±1.20 | 0.15 |
| White blood cells (x1000 cells/mm<sup>3</sup>) | 3.20±0.60 | 4.34±0.94 | 0.07 |
| Neutrophils (%) | 66.30±9.70 | 75.00±10.00 | 0.14 |
| Lymphocytes (%) | 27.00±10.00 | 17.00±7.00 | 0.10 |
| Eosinophils (%) | 0.60±0.40 | 1.00±1.00 | 0.84 |
| AST (U/L) | 32.00±20.00 | 58.50±18.00 | 0.06 |
| ALT (U/L) | 29.00±14.00 | 43.15±11.45 | 0.11 |

CI – Confidence level

The data for microRNA expression within 4 days from fever onset is normally distributed at 95% confidence interval. hsa-miR-150-5p expression showed significant (P<0.05) upregulation in DHF (n=20) patients compared to DF (n=20) patients within 4 days, day 3 (n_DF_=6, n_DHF_=12) and within 3 days (n_DF_=8, n_DHF_=15) from fever onset before the patients presented with symptoms of severe illness. Samples collected on day 2 (n_DF_=2, n_DHF_ =3) and day 4 (n_DF_=11, n_DHF_=5) from fever onset failed to show differential expression (Fig 1a, Table S4). This may be due to limited number of samples available for expression analysis collected within 2 days from fever onset. While both males and females showed similar upregulation in expression of hsa-miR-150-5p in DHF patients within 4 days from fever onset, this upregulation was not statistically significant (Fig 1b, Table S5). This may be due to the relatively low number of female samples available for the analysis. The rest of the microRNA under investigation also did not show significant differential expression in DHF patients over DF patients within 4 days from fever onset (Fig 1a).

**Fig 1.**
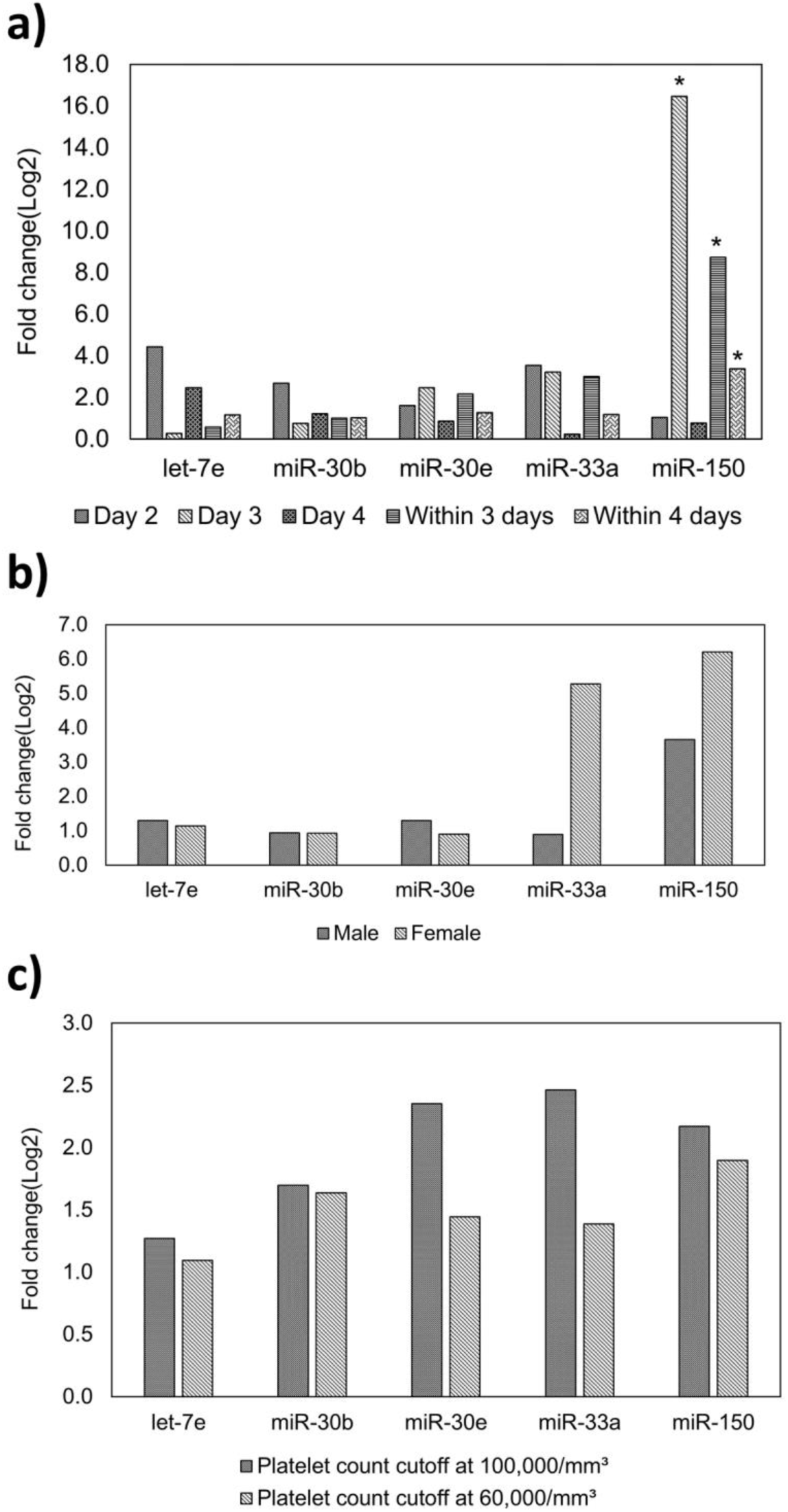
Fold change of expression of hsa-let-7e (let-7e), hsa-miR-30b-5p (miR-30b), hsa-miR-30e-3p (miR-30e), hsa-miR-33a (miR-33a) and hsa-miR-150-5p (miR-150) in PBC samples from DF and DHF patients. (**a)** recruited on, day 2 (n_DF_=2, n_DHF_ =3), day 3 (n_DF_=6, n_DHF_=12), day 4 (n_DF_=11, n_DHF_=5), within 3 days (n_DF_=8, n_DHF_=15) and within 4 days (n_DF_=20, n_DHF_=20) from fever onset (**b)** male (n_DF_=14, n_DHF_=17) and female (n_DF_=6, n_DHF_=3) recruited within 4 days from fever onset (**c)** recruited within 4 days from fever onset with platelet count > 100 × 10^3^ cells/ mm^3^ (n=9) compared to those < 100 × 10^3^ cells/ mm^3^ (n=31) and platelet count > 60,000 cells/mm^3^ (n=14) compared to those < 60,000 cells/ mm^3^ (n=26) during the course of infection presented as log values to the base 2 based on ΔΔCq values against hsa-miR-16-5p and hsa-miR-103a-3p, where a fold change >1.5 was considered as up regulation and < 0.5 considered as down regulation. * P<0.05 based on ΔCq ± SEM using independent t - test.

According to the WHO guidelines, a rapid progressive drop in platelet count below 100,000/mm^3^ and a rising [2]. hematocrit level above baseline (> 52% for male and >48% for female) is considered a major risk factor for manifestation of severe dengue symptoms. Platelet count < 60,000 /mm^3^ is also considered as a better cut off for severe dengue marker [33]. The patients recruited for the study did not have hematocrit levels above the baseline. (Table S3). Therefore, we evaluated the differential expression of microRNA between the patients with platelet count < 100,000/mm^3^ during the course of hospitalization (n=31) and those who maintained a platelet count > 100,000/mm^3^ during the course of hospitalization (n=9) as well as platelet count < 60,000 /mm^3^ (thrombocytopenia) (n=26) and >60,000 /mm^3^ (n=14) during the course infection. hsa-miR-30b-5p, hsa-miR-30e-3p, hsa-miR-33a, and hsa-miR-150-5p showed upregulation (>1.5) of expression in patients with platelet count <100,000/mm^3^ (thrombocytopenia) compared to those with platelet count > 100,000/mm^3^ while hsa-miR-30b-5p and hsa-miR-150-5p showed upregulation (>1.5) in patients with platelet count < 60,000 /mm^3^ compared to those with platelet count > 60,000 /mm^3^ during the course of infection. However, these differences were not significant. (Fig 1c, Table S6). Therefore, the microRNA under investigation did not serve as markers of severity of infection as marked by thrombocytopenia during the acute phase of infection.

Logistic regression analysis for miR-150 expression based on ΔCq values was found to be predictive of DHF within 4 days from fever on set with odds ratio of 0.76 (95%, CI;0.58-0.99, P=0.04) and within 3 days with odds ratio of 0.52 (95%, CI;0.29-0.93, P=0.03) (Table S7). The area under the receiver operating characteristic curve (AUC) for miR-150 expression is 0.70 (sensitivity 0.65, specificity 0.75) within 4 days at ΔCq of 7.68 (P<0.05), 0.85 (sensitivity 0.80, specificity 0.88) within 3 days at ΔCq of 7.54 (P<0.05) and 0.92 (sensitivity 1.00, specificity 0.50) for day 3 from fever onset at ΔCq of 9.25 (P<0.05) indicating that miR-150 may serve as an early marker for development of severe manifestations of dengue within 4 days from fever onset (Table S8, Fig S1).

### Differential expression analysis of putative target mRNA of the selected microRNA in Dengue patients within four days from fever onset

MicroRNA suppress or inhibit the expression of putative target genes. As such, the upregulation of microRNA expression may lead to the downregulation of the putative target gene expression. Therefore, relative expression of putative target genes of the microRNA under investigation, EZH2, ABCA1, DNMT3A and RIP140 were evaluated against the expression of GAPDH as a reference gene in DF (n=20) patients who tested positive for NS1 antigen and those who later developed DHF (n=20) within four days from fever onset.

All the data for the target gene expression were normally distributed. EZH2 expression showed significant down regulation (P<0.05) in DHF patients within 4 days from fever onset compared to that in DF patients. Evaluation of differential expression in DF and DHF samples from patients recruited day 3 (n_DF_=6, n_DHF_=12), day 4 (n_DF_=11, n_DHF_=5) and within 3 days (n_DF_=8, n_DHF_=15) from fever onset also showed a downregulation in EZH2 expression. However, the downregulation in expression was only significant (P<0.05) in samples from patients recruited on day 4 from fever onset (Fig 2a, Table S9). These findings are consistent with upregulation of miR-150 in PBC of DHF patients compared to DF patients at the early stages.

**Fig 2.**
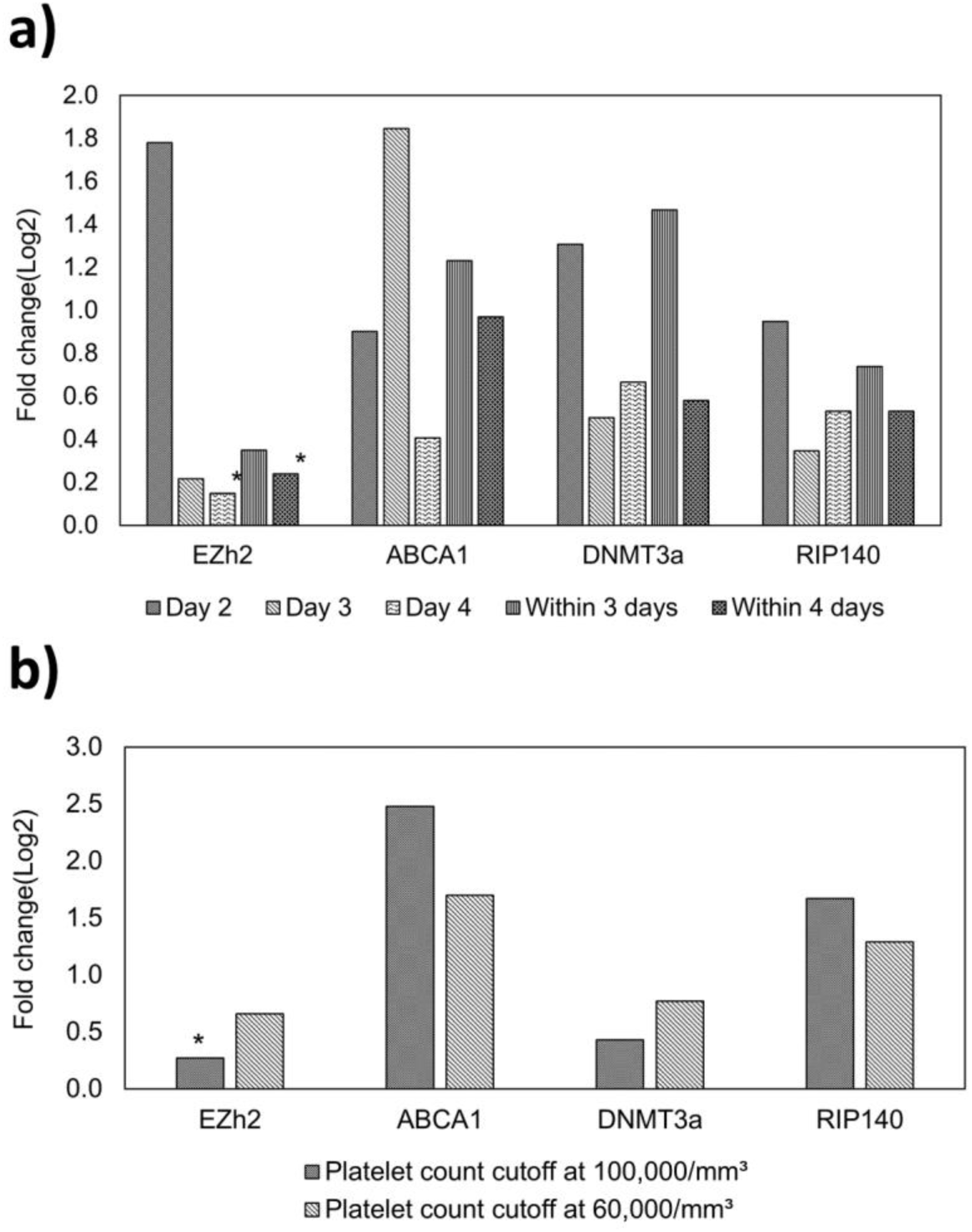
Fold change of microRNA target gene expression in PBC between DF and DHF patients. (**a)** recruited on, day 2 (nDF=2, nDHF =3), day 3 (nDF=6, nDHF=12), day 4 (nDF=11, nDHF=5), within 3 days (nDF=8, nDHF=15) and within 4 days (nDF=20, nDHF=20) from fever onset (**b)** recruited within 4 days from fever onset with platelet count > 100 × 10^3^ cells/ mm^3^ (n=9) compared to those < 100 × 10^3^ cells/ mm^3^ (n=31) and platelet count > 60 × 10^3^ cells/mm^3^ (n=14) compared to those < 60 × 10^3^ cells/ mm^3^ (n=26) during the course of infection presented as log values to the base 2 based on ΔΔCq values against GAPDH, where a fold change >1.5 was considered as upregulation and < 0.5 considered as downregulation. * P<0.05 based on ΔCq ± SEM using independent t - test.

Logistic regression analysis for EZH2 expression on day 4 is found to be predictive for DHF with odds ratio of 4.23 at 95% CI; 0.93 – 19.34, AUC of 0.91 with sensitivity of 0.60 and specificity of 0.82 at ΔCq of 3.44 and odds ratio for expression within 4 days from fever is 1.60 at 95% CI; 1.12 – 2.31 (P=0.01), AUC of 0.76 with sensitivity of 0.85 and specificity of 0.45 at ΔCq of 2.04 (P=0.01) (Tables S10, S11 and Fig S2). ABCA1, DNMT3A and RIP140 failed to show differential expression within 4 days from fever onset among the DF and DHF samples. ABCA1 showed downregulation of expression only among patient samples collected on day 4 from fever onset.

We also evaluated the expression of these target genes of microRNA within 4 days from fever onset, among patients with platelet count <100,000/mm^3^ (n=31) and platelet count < 60,000/mm^3^ (n=26) as indicative of severe manifestation of dengue based on thrombocytopenia compared to those with platelet count >100,000/mm^3^ (n=9) and > 60,000/mm^3^ (n=14) during the course of infection. EZH2 showed significant (P<0.05) downregulation within 4 days from fever onset in patients with platelet count <100,000/mm^3^ (n=31) compared to those with platelet count > 100,000/mm^3^ (n=9) during the course of infection suggesting a role for EZH2 in severe manifestation of dengue as marked by thrombocytopenia at a platelet count cutoff of 100,000/mm^3^ (Fig 2b, Table S12). While EZH2 expression is also downregulated among patients at a platelet count cutoff of 60,000, none of the putative target genes show significant differential expression among the patients with platelet count <60,000/mm^3^ and >60,000/mm^3^ (Fig 2b, Table S12).

Odds ratio for development of severe dengue marked by platelet count <100,000/mm^3^ based on logistic regression analysis for EZH2 expression within 4 days from fever onset is 1.46 (95**%** CI;1.00**-** 2.12, P**=**0.05), AUC 0.82 with sensitivity of 0.84 and specificity of 0.60 (P=0.02) at ΔCq of 2.04 suggesting that EZH2 may serve as an early marker of severe dengue marked by development of thrombocytopenia during the course of infection (Tables S10 and S11).

## Discussion

miR-150 is significantly upregulated (p<0.05) in DHF patients on day 3, within 3 days and within 4 days from fever onset. Therefore, upregulation of miR-150 and the indirect effects of miR-150 on EZH2 may have contributed to the downregulation of EZH2 expression. Although hsa-miR-150-5p showed significant upregulation (P<0.05) in DHF samples within four days from fever onset, samples collected on day 4 did not show significant differential expression among the DF and DHF patients. Therefore, day 3 may be a better cutoff for utility of miR-150 expression as a prognsitic marker for severity of dengue infection. Evaluating differential microRNA expression in a larger cohort of patient samples is needed to evaluate the fluctuations in the level of microRNA from fever onset during the early stages of infection. Calculated sample sizes are (n_DF_ =15218, n_DHF_ =22828) on day 2, (n_DF_ =5, n_DHF_ =9) on day 3, (n_DF_ =1479, n_DHF_ =665) on day 4, (n_DF_ =7, n_DHF_ =13) within 3 days, (n_DF_ =35, n_DHF_ =35) within 4 days. Therefore, based on the calculated sample sizes, sufficient number of samples have been analyzed to determine the ability of miR150 to serve as an early biomarker of severity of infection in dengue patients on day 3.

Calculated sample size with 95% confidence interval and 80% power for biomarker validity of EZH2 is 17 each for within 4 days from fever onset, n_DF_ =10 and n_DHF_ = 4 for 4 days and n_DF_ =9 and n_DHF_ =12 for 3 days from fever onset indicating sufficient number of samples have been analyzed to determine the ability of EZH2 to serve as an early biomarker of severity of infection in dengue patients. Significant downregulation (P<0.05) of EZH2 within 4 days from fever onset may also serve as an early biomarker for severe dengue marked by thrombocytopenia (<100,000/mm^3^) during the course of infection. The calculated sample sizes for EZH2 to serve as a biomarker for severity of infection based on platelet count are n>100,000/mm^3^ = 17, n<100,000/mm^3^ = 57, n>60,000/mm^3^ = 219, n<60,000/mm^3^ = 407, suggesting the need for validation in a larger cohort of samples. Further studies in cultured cells may be needed to evaluate the role of miR150 on EZH2 expression during severe dengue virus infections.

Regulation of miR-150 and EZH2 expression has also been linked to inducible Nitric Oxide Synthase (iNOS or NOS2) activity where iNOS is indirectly regulated by miR-150. A study showed, that infection of Mycobacterium bovis bacillus Calmette-Guérin (BCG) regulates the epigenetic changes at class II transactivator (CIITA) promoter via induction of expression of iNOS/NO and miR-150 through the recruitment of the transcription factor. According to our studies, differential expression analysis of iNOS in DF and DHF patient samples collected on day 2, 3, 4, within 3 days and within 4 days from fever onset showed downregulation in DHF patients as early as day 2 from fever onset and remained downregulated [34].

let-7e, which has been reported to directly regulate EZH2 expression is only upregulated in DHF patients on day 2 and day 4 from fever onset while the expression is downregulated on day 3. Calculated sample sizes for the role of the microRNA under investigation as early biomarkers of severity of infection based on the data from 20 DF and and 20 DHF samples within 4 days from fever onset are given in Tables S13-S15. Differential expression of EZH2 as a result of upregulation of let-7e expression was implicated in several viral infections including DENV2 infected PBMCs [35]. Inhibition of EZH2 expression repressed the herpes simplex viral gene expression [36]. Therefore, further studies in dengue virus infected cultured cells may be needed to evaluate the role of let-7e on the differential expression of EZH2 during severe dengue virus infection.

Epigenetic modifications are hereditary DNA or histone modifications that do not change the DNA sequence and affect transcriptional regulation. EZH2 is an epigenetic modifier that can catalyze the methylation of histone H3 protein. miR-150 plays a critical role in epigenetic modifications involving regulating EZH2 expression at CIITA promoter region during the M. bovis BCG infection [27]. Therefore, epigenetic modifications may play a role in severe manifestation of dengue during secondary infections. Differential expression of epigenetic modifiers such as EZH2 in patients marked by plasma leakage and**/**or thrombocytopenia suggests that there is a role for epigenetic regulation in severe manifestations of dengue. The polycomb group protein, EZH2 and DNMT3A are both involved in epigenetic regulation via DNA methylation. Binding of DNMTs to several EZH2-repressed genes depends on the presence of EZH2. EZH2 serves as a recruitment platform for DNA methyltransferases [37]. Since both EZH2 and DNMT3A participate in epigenetic modification mediated transcriptional regulation, differential expression analysis in virus infected cultured cells is needed to further evaluate the role of DNMT3A in severe dengue. Expression of ABCA1 and RIP140 implicated in lipid metabolism, a mechanism which has been shown to play a role in viral infections on the other hand did not significantly change in patients with severe dengue (Fig 2a) [32]. While several putative target genes of miR150 should be validated in a larger patient cohort based on the calcuated sample sizes (Tables S16 and S17), our studies have shown that differential expression of microRNA miR-150-5p and the putative target gene EZH2 may serve as reliable biomarkers of disease severity during early stages of dengue infection when differential diagnosis or prognosis is not possible based on the clinical evidence.

## Data Availability

The data used to support the findings of this study are included within the article and within the supplementary information files.

## Supporting information

**Table S1. Primers for microRNA target genes for qRT-PCR analysis, NCBI accession code and previous publications (DOCX)**

**Table S2. Primers for microRNA genes for qRT-PCR analysis, NCBI accession code and previous publications (DOCX)**

**Table S3. AST, ALT and hematocrit levels of patients from admission to discharge (DOCX)**

**Table S4. Relative expression of microRNA presented as fold change based on ΔΔCq values against hsa-miR-16-5p and hsa-miR-103a-3p (Log**_**2**_**) and** Δ**Cq at 95% confidence intervals (CI) (DOCX)**

**Table S5. Relative expression of microRNA in male and female patients within 4 days from fever onset presented as fold change based on ΔΔCq values against hsa-miR-16-5p and hsa-miR-103a-3p (Log**_**2**_**) and** Δ**Cq at 95% confidence intervals (CI) (DOCX)**

**Table S6. Relative expression of microRNA at platelet count cut off 100**,**000/mm**^**3**^ **and 60**,**000/mm**^**3**^ **within 4 days from fever onset presented as fold change based on ΔΔCq values against hsa-miR-16-5p and hsa-miR-103a-3p (Log**_**2**_**) and** Δ**Cq at 95% confidence intervals (CI) (DOCX)**

**Table S7. Odds ratio for microRNA based on Logistic regression analysis (95%, CI) (DOCX)**

**Table S8. Receiver operating characteristic curve for microRNA (DOCX)**

**Table S9. Relative expression of putative target genes of microRNA presented as fold change based on ΔΔCq values against GAPDH (Log**_**2**_**) and** Δ**Cq at 95% confidence intervals (CI) (DOCX)**

**Table S10. Odds ratio for putative microRNA target genes based on Logistic regression analysis (95%, CI) (DOCX)**

**Table S11. Receiver operating characteristic curve for putative microRNA target genes (DOCX)**

**Table S12. Relative expression of putative target genes of microRNA at platelet count cut off 100**,**000/mm**^**3**^ **and 60**,**000/mm**^**3**^ **within 4 days from fever onset presented fold change based on ΔΔCq values against GAPDH (Log**_**2**_**) and** Δ**Cq at 95% confidence intervals (CI) (DOCX)**

**Table S13. Calculated sample sizes for microRNA of DF and DHF groups at 95% confidence level and power of 80 (DOCX)**

**Table S14. Calculated sample sizes for microRNA for male and female DF and DHF groups at 95% confidence level and power of 80. Samples collected within 4 days from fever onset (DOCX)**

**Table S15. Calculated sample sizes for microRNA at platelet count cut off 100**,**000/mm**^**3**^ **and 60**,**000/mm**^**3**^, **within 4 days from fever onset at 95% confidence level and power of 80 (Platelet count determined during the course of hospitalization**) **(DOCX)**

**Table S16. Calculated sample sizes for putative target genes of DF and DHF groups at 95% confidence level and power of 80 (DOCX)**

**Table S17. Calculated sample sizes for putative target genes at platelet count cut off 100**,**000/mm**^**3**^ **and 60**,**000/mm**^**3**^, **within 4 days from fever onset at 95% confidence level and power of 80 (Platelet count determined during the course of hospitalization**) **(DOCX)**

**Figure S1. ROC curves for miR150 on (a) day 3, (b) within 3 (c) within 4 days from fever onset (DOCX)**

**Figure S2. ROC curves for EZH2 on (a) day 4, and (b) within 4 days from fever onset (DOCX)**

## Funding

This work was supported by University of Kelaniya (RP/03/SR/02/06/02/2016); ECWS Fellowship (4500384850) and National Science Foundation, Sri Lanka (RG/2015/BT/02).

## Conflicts of interest

The authors declare to have no conflicts of interest

## Author Contributions

Conceptualization: Nimanthi Jayathilaka

Formal analysis: Nimanthi Jayathilaka

Funding acquisition: Nimanthi Jayathilaka

Investigation: Harsha Hapugaswatta, Pubudu Amarasena, Ranjan Premaratna

Methods: Nimanthi Jayathilaka

Project administration: Nimanthi Jayathilaka, Kapila N. Seneviratne

Resources: Ranjan Premaratna, Pubudu Amarasena

Supervision: Nimanthi Jayathilaka, Kapila N. Seneviratne

Writing - original draft: Nimanthi Jayathilaka, Kapila N. Seneviratne

Writing – review and editing: Nimanthi Jayathilaka, Kapila N. Seneviratne

## Notes

### Competing Interest Statement

The authors have declared no competing interest.

### Author Declarations

All relevant ethical guidelines have been followed and any necessary IRB and/or ethics committee approvals have been obtained.

Any clinical trials involved have been registered with an ICMJE-approved registry such as ClinicalTrials.gov and the trial ID is included in the manuscript.

